# Initial Feasibility and Clinical Implementation of Daily MR-guided Adaptive Head and Neck Cancer Radiotherapy on a 1.5T MR-Linac System: Prospective R-IDEAL 2a/2b Systematic Clinical Evaluation of Technical Innovation

**DOI:** 10.1101/2020.06.22.20137554

**Authors:** Brigid A. McDonald, Sastry Vedam, Jinzhong Yang, Jihong Wang, Pamela Castillo, Belinda Lee, Angela Sobremonte, Yao Ding, Abdallah S.R. Mohamed, Peter Balter, Neil Hughes, Daniela Thorwarth, Marcel Nachbar, Marielle E.P. Philippens, Chris H.J. Terhaard, Daniel Zips, Simon Böke, Musaddiq J. Awan, John Christodouleas, Clifton D. Fuller On Behalf of the MR-Linac Consortium Head and Neck Tumor Site Group and the Joint Head and Neck Radiotherapy-MRI Development Cooperative

## Abstract

**Introduction:** This prospective study is the first report of daily adaptive radiotherapy (ART) for head & neck cancers (HNC) using a 1.5T MR-linac, with particular focus on safety & feasibility and dosimetric results of an on-line rigid registration-based adapt-to-position (ATP) workflow.

**Methods:** Ten HNC patients received daily ART on a 1.5T/7MV MR-linac, six using ATP only and four using ATP with one off-line adapt-to-shape re-plan. Setup variability with custom immobilization masks was assessed by calculating the average systematic error (M), standard deviation of the systematic error (∑), and standard deviation of the random error (σ) of the isocenter shifts. Quality assurance was performed with a cylindrical diode array using 3%/3mm γ criteria. Adaptive treatment plans were summed for each patient to compare delivered dose with planned dose from the reference plan. The impact of dosimetric variability between adaptive fractions on the summation plan doses was assessed by tracking the number of optimization constraint violations at each individual fraction.

**Results:** The random errors (mm) for the x, y, and z isocenter shifts, respectively, were M = − 0.3, 0.7, 0.1; ∑ = 3.3, 2.6, 1.4; and σ = 1.7, 2.9, 1.0. The median γ pass rate was 99.9% (range: 90.9%-100%). The differences between the reference and summation plan doses were within [-0.61%, 1.78%] for the CTV and [-11.74%, 8.11%] for organs at risk (OARs), though percent increases in OAR dose above 2% only occurred in three cases, each for a single OAR. All cases had at least two fractions with one or more constraint violations. However, in nearly all instances, constraints were still met in the summation plan despite multiple single-fraction violations.

**Conclusion:** Daily ART on a 1.5T MR-linac using an on-line ATP workflow is safe and clinically feasible for HNC and results in delivered doses consistent with planned doses.

## 1. Introduction

High-field magnetic resonance imaging (MRI)-guided radiation therapy (RT) has become a clinical reality with the commercial implementation and health-system accreditation of a hybrid 1.5T/7MV MRI/linear accelerator (MR-linac) [1–3]. The enhanced soft tissue contrast of the MR-linac allows visualization of non-bony target volumes and organs at risk (OARs) during treatment setup, enabling daily on-line treatment plan reoptimization for adaptive radiotherapy (ART) [3–5]. First clinical experiences with the MR-linac have been previously reported for oligometastatic, pelvic, and breast tumors [6–9], but utilization of the device has since expanded to a number of other tumor sites, including head and neck cancer (HNC).

Rather than *ad hoc* technology development and implementation via unstructured, non-sequential processes, the MR-Linac Consortium [10] has developed the R-IDEAL conceptual framework of systematic technical and clinical reporting processes for clinical radiotherapy applications [11]. Furthermore, to our knowledge, there are no reports of high-field MR-guided daily ART for HNC on a hybrid MR-linac system. Thus, pursuant to this commitment, we sought to undertake an R-IDEAL Stage 2a (“technical optimization of the innovation for treatment delivery”)/2b (“proof of early clinical effectiveness and safety of the innovation”) study with the following aims:

1) Describe an adapt-to-position (ATP) workflow for daily ART using conventional fractionation and ensuring preservation of pre-therapy dose constraints for target volumes and OARs; 2) Demonstrate the capacity of the 1.5T MR-linac system to deliver safe and clinically acceptable ART plans for HNC; 3) Characterize performance based on treatment times, quality assurance (QA) results, setup corrections, cumulative doses, and dosimetric variability between adaptive plans.

## 2. Methods

### 2.1 MR-Linac Clinical Workflow for Head and Neck

The clinical workflow described in this paper was developed as a multi-institutional effort through the MR-Linac Consortium’s Head and Neck Tumor Site Group. Although initial implementation at a single site (MD Anderson Cancer Center; Houston, TX) is described, this workflow is based on guidelines iterated through the Tumor Site Group prior to large-scale implementation (Appendix A).

#### 2.1.1 Pre-treatment Workflow

##### Patient Selection

HNC patients are deemed eligible for treatment on the 1.5T/7MV MR-linac (Unity; Elekta AB; Stockholm, Sweden) if the following criteria are met: Patients must have intact tumor or an MR-visible reference OAR (e.g. skull base) and planning target volume (PTV) length <20 cm in the super-inferior direction (due to field size constraints [12]). Patients with claustrophobia, contraindications to MRI (metal objects, pacemakers, etc.), and/or compromised airway are excluded. Because the MR-linac is located in the out-patient radiation treatment center at our institution, the patient must be ambulatory. If the patient is admitted to the in-patient facilities at any time during treatment, he or she must be treated with a back-up plan on a conventional linac at the in-patient center. The full Patient Evaluation Form and MRI Safety Screening Form are given in Appendices B and C.

##### Simulation Protocol

All patients receive both CT and MR simulations on the same day and with the same immobilization devices to ensure that the CT and MR images may be rigidly registered [13]. The CT simulation is used for planning for both the MR-linac treatment and the back-up plan on a conventional linac. A tabletop overlay is used on the CT scanner to reproduce the actual couch top in the MR-linac treatment room. Placement of the index bars along the couch establishes the location of patient immobilization aids and the actual tumor volume with respect to the isocenter on the MR-linac. The tabletop overlay is also equipped with a hoop-shaped device representing the maximum clearance of the MR-linac bore to ensure that the patient and all immobilization aids will be able to fit into the MR-linac. The patient is immobilized with a custom thermoplastic immobilization mask and cushion (Klarity Medical Products; Heath, OH) (Figure 1).

**Figure 1:**
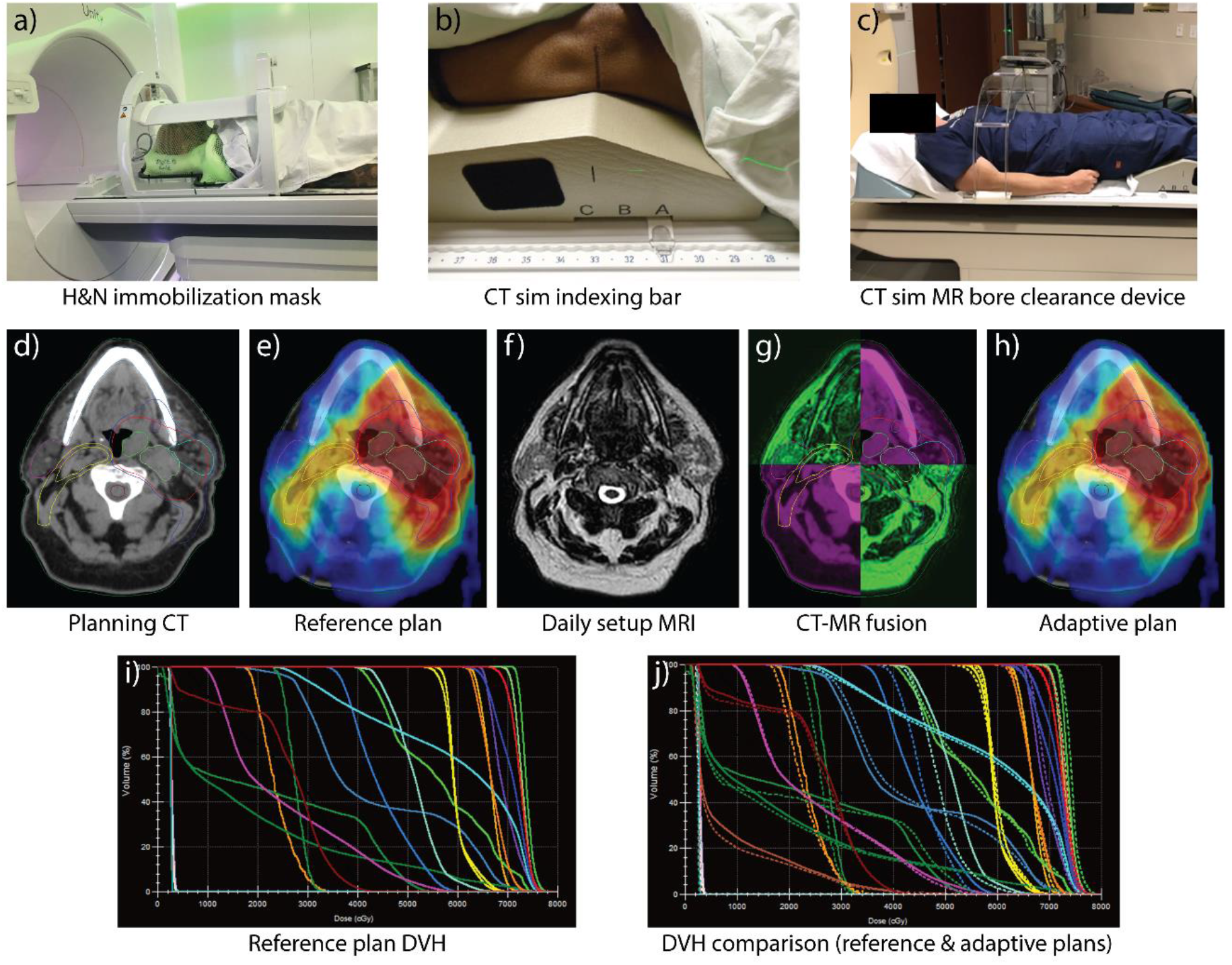
The MR-linac simulation and on-line adaptive replanning workflow. a) Custom head & neck immobilization mask with the coil on the MR-linac; b) indexing bar on the CT simulation MR-linac table overlay; c) CT sim hoop device used to test clearance in MR-linac bore; d) planning CT with contours; e) reference treatment plan on planning CT; f) daily setup MRI from MR-linac (2-minute T2 protocol); g) rigid registration of planning CT and setup MRI; h) adaptive plan using ATP workflow; i) dose volume histogram (DVH) of reference plan; j) DVH comparison between reference and adaptive plans.

The MR simulation protocol includes two standard vendor-provided non-contrast T2-weighted 3D MRI sequences (2-minute and 6-minute scans; scan protocol parameters provided in Appendix D) and may include additional specialized sequences (diffusion-weighted imaging (DWI), 2D balanced fast field echo (bFFE) in three orthogonal directions for motion estimation, etc.). The physician determines at the time of MR simulation whether the 2-minute T2-weighted scan is adequate for daily setup and plan reoptimization; if the tumor or reference OAR is not clearly visible, the 6-minute scan is used for daily imaging.

##### Treatment Planning and Pre-Treatment Preparation

The CT and T2-weighted MR simulation images are fused for contouring of the target volume(s) and OARs by the physician and dosimetrist, respectively. Treatment planning is done in Monaco (version 5.4; Elekta AB), which employs a Monte Carlo-based dose calculation engine. All MR-linac plans use step-and-shoot intensity-modulated radiation therapy (IMRT). An initial reference plan is created by a dosimetrist based on the target prescription dose(s) and OAR constraints (Appendix E) provided in the physician’s planning directive. The optimized plan is reviewed by the physician and physicist. Upon approval, the treatment plan is exported to the radiotherapy treatment data management system (Mosaiq; Elekta AB), where a Plan of Care is created for the patient. The Plan of Care specifies the exact MRI exam card(s) to be loaded during each treatment session.

#### 2.1.2 Online Clinical Workflow

The Unity on-line adaptive workflow has been previously described by Winkel *et al*. [3]. At our institution, a T2-weighted MRI is used for daily setup verification and plan adaptation for HNC patients. The 6-minute scan was used for the first three patients to be conservative, but the 2-minute scan was used for all subsequent patients because the target or reference anatomy could be adequately visualized. The daily setup image is then fused with the reference plan image.

Two on-line adaptive workflows are possible with the Unity system: adapt-to-position (ATP) and adapt-to-shape (ATS). ATP is a virtual isocenter shift of the reference plan based on rigid registration with either a dose recalculation or plan reoptimization. ATS involves deformable image registration to propagate contours onto the daily setup image followed by a full plan reoptimization on the current anatomy [3]. The ATP workflow was used for on-line plan adaptation for six patients, and a combination of on-line ATP and off-line ATS was used for four patients (maximum one ATS during treatment course) to account for soft tissue deformation.

The following acceptance criteria are used for on-line plan review and to determine when an ATS is required: After fusion of the setup and reference plan images, if the isocenter shift is <5 mm in all directions and planning constraints are satisfied, the ATP workflow can proceed. If all shifts are <5 mm but any one dosimetric criterion is not satisfied for three consecutive fractions, the physician must be consulted prior to treatment and an off-line ATS is considered. If any shift is >5 mm, the patient must be repositioned; if any shift is still >5 mm (most likely due to patient weight loss), the physician must approve the plan prior to treatment, and an off-line ATS will be performed.

On-line ATS is challenging for HNC due to the large number of structures used in plan optimization. To improve the efficiency of the on-line workflow, ATS was only performed off-line to create a new reference plan for use in the subsequent days’ on-line ATP. When an ATS is performed, a 6-minute T2-weighted MRI is acquired during the previous fraction as the reference image for the ATS plan because the 2-minute scan is not adequate for target delineation. In the current study, the goal of treatment plan reoptimization was to ensure the delivery of pre-therapy planned doses to target volumes and OARs, referred to as an ART_ex_aequo_^1^approach in the nomenclature defined by Heukelom and Fuller [14]. Thus, in any off-line ATS’s, the target structures were registered rigidly rather than deformably and were not modified in any capacity; only OAR contours were updated to reflect anatomical changes.

Four methods are available for plan recalculation/reoptimization: “original segments,” “adapt segments,” “optimize weights,” and “optimize weights and shapes” [3]. In our ATP workflow, we use “adapt segments” when the isocenter shift is <1 mm in all directions. Otherwise, we use “optimize weights” first then “optimize weights and shapes” if the plan does not satisfy dosimetric criteria. For ATS, we use “optimize weights and shapes.”

During on-line plan adaptation, motion monitoring with a bFFE sequence is turned on to ensure that the patient has not deviated from the original position. Once the adaptive plan is created, it must be approved by the physicist and physician. Next, a secondary monitor unit (MU) calculation is performed using RadCalc (LAP/LifeLine Software, Inc; Austin, TX) for beam-by-beam validation of the MUs in the adaptive plan. Upon final approval, the treatment is delivered. IMRT QA is performed after the treatment using a cylindrical diode array.

### 2.2 Patient Cohort and Informed Consent

This prospective study included the first ten HNC patients at our institution who satisfied the following criteria: treated with multiple IMRT fractions on the MR-linac, enrolled in the MOMENTUM observational clinical trial (NCT04075305; institutional IRB PA18-0341), and not enrolled in any other clinical trials involving non-conventional dosing or fractionations schemes. These patients provided written informed consent for their images, treatment plans, and clinical data to be used.

### 2.3 Treatment Times, IMRT Quality Assurance, and Setup Variability

Treatment times were calculated from the time stamps in the record & verify system (Mosaiq). Three times are reported: 1) setup and plan reoptimization (the time between the patient’s arrival at the MR-linac and the delivery of the first beam); 2) beam delivery (the time between delivery of the first and last beam); and 3) total treatment time.

IMRT QA was performed for every adaptive plan following treatment. An MR-compatible cylindrical diode array (ArcCHECK MR; Sun Nuclear Corporation; Melbourne, FL) was used with 3% dose/3mm distance to agreement γ criteria. A γ value of 90% or more was considered passing.

Daily setup variability was quantified by recording the isocenter shifts along the x (left/right), y (superior/inferior), and z (anterior/posterior) axes at each adaptive treatment delivered on the MR-linac (253 fractions among 10 patients). Systematic and random errors in each direction were calculated using Van Herk’s method [15], such that M represents the mean systematic error of all isocenter shifts for all patients, ∑ represents the standard deviation of the systematic error, and σ represents the standard deviation of the random error.

### 2.4 Comparison of Reference Plan and Daily Adaptive Plans

In order to validate the daily plan quality generated by the daily ATP workflow, the initial reference plan was compared to the accumulated dose of the daily adaptive plans (summation plan) for each patient. In the Unity ATP workflow, all treatment plans generated from the same reference plan have their dose distributions calculated on the reference image set (i.e. the treatment plans and doses are already in the same frame of reference). This enables rigid dose summation of the ATP plans, whereby all daily adaptive plan doses are scaled to a single fraction then added voxel-by-voxel. In this study, daily adaptive plans were summed using rigid dose summation on the reference image set to create the summation plan. When ATS was performed, the ATS reference image was rigidly registered to the initial reference image prior to plan summation.

Plans were evaluated using the actual values of the quantities used as dosimetric constraints for each patient, such as maximum dose (D_max_) to the spinal cord or mean dose (D_mean_) to the parotid glands (Appendix E). In cases where there were multiple prescription dose levels to different target volumes, only the highest dose was considered for analysis. The summation plan and reference plan for each patient were compared by calculating the percent difference of each plan quality metric:

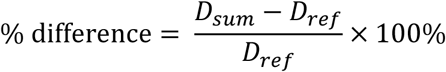

In cases where the backup plan was delivered on a conventional linac for one or more fractions, only the MR-linac plans were considered for dosimetric analysis by scaling the reference plan to the number of fractions delivered on the MR-linac prior to taking the percent difference. The cumulative dose at each fraction was calculated for one representative case and compared to the expected dose (the cumulative dose if the initial reference plan had been delivered at each fraction). The number of times an adaptive plan violated a dosimetric constraint was also tracked for each patient.

## 3. Results

### 3.1 Patient Cohort

The patient cohort included eight males and two females with median age 62 (range: 39–80). The most common treatment sites were larynx (n=3) and oropharynx (n=3). A total of 253 adaptive treatments were delivered on the MR-linac among the ten patients. Seven patients received at least one treatment with their backup plan on a conventional linac due to machine downtime and/or admittance to in-patient facilities. All patients were treated with the ATP on-line workflow, with four patients receiving a mid-treatment off-line ATS re-plan due to anatomical changes. Full treatment information is provided in Table 1.

**Table 1:**
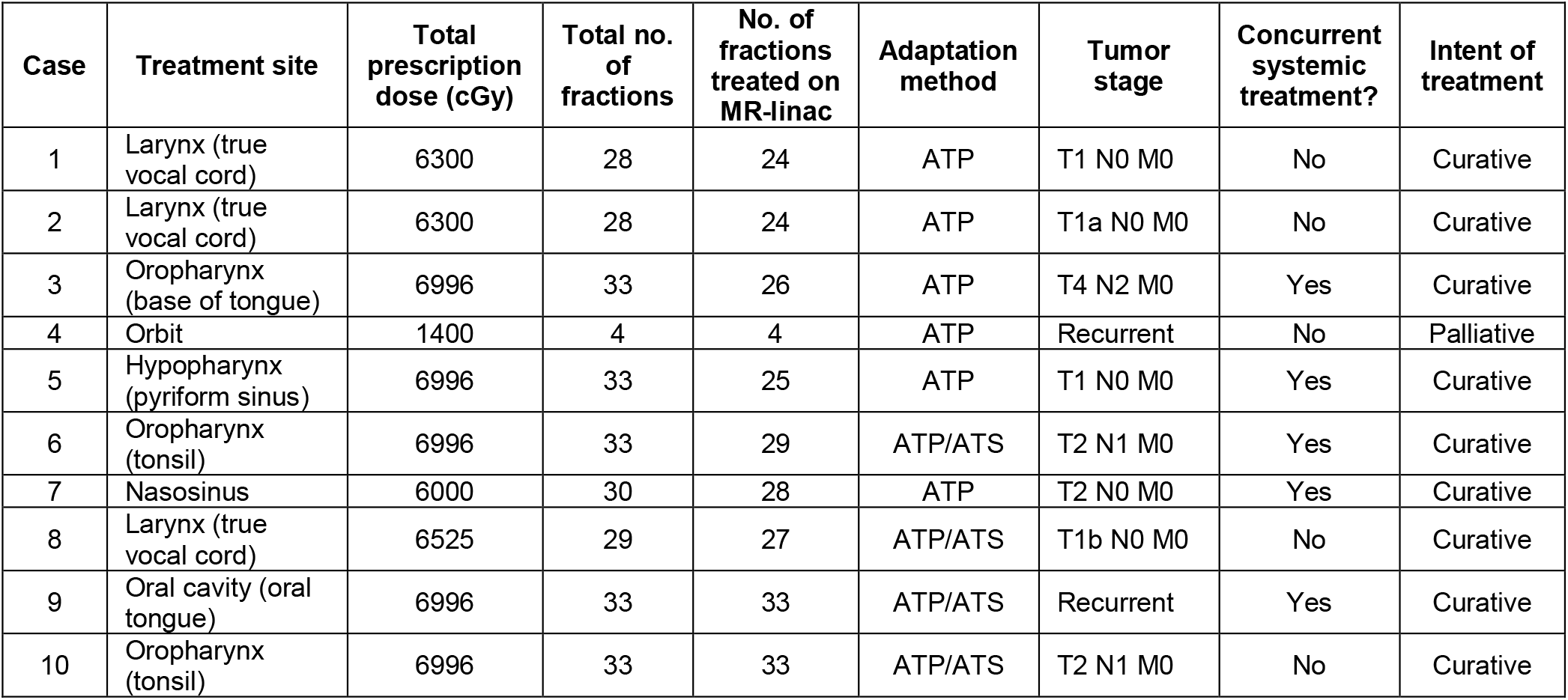
Patient treatment information for the 10 cases.

### 3.2 Treatment Times, IMRT Quality Assurance, and Setup Variability

The minimum, lower quartile, median, upper quartile, and maximum treatment times (minutes) were 20, 27, 31, 38, 71 for setup and plan reoptimization; 7, 13, 13, 16, 28 for beam delivery; and 31, 41, 46, 53, 85 for total treatment time, respectively. 91% of total treatment times were under 60 minutes.

All adaptive plans in this study passed IMRT QA with a γ value greater than 90% (Appendix F). The minimum, lower quartile, median, upper quartile, and maximum γ values were 90.9%, 99.4%, 99.9%, 100.0%, and 100.0%, respectively. 42.5% of all plans had a γ value of 100%, indicating excellent agreement between the expected and delivered dose distributions overall. The absolute value mean systematic error of the isocenter shifts was ≤ 0.7 mm in all 3 orthogonal directions, and the systematic and random errors were ≤ 3.3 mm (Table 2). The maximum absolute value isocenter shifts in any single fraction were 8.0, 19.7, and 7.3 mm in the x, y, and z directions, respectively. In the y-direction, the shift exceeded 1 cm only four times (Appendix F).

**Table 2:** Average systematic error (M), standard deviation of the systematic error (∑), and standard deviation of the random error (σ) of the x, y, and z isocenter shifts during daily setup of 10 patients (253 adaptive fractions on the MR-linac).

| Axis | $M$ (mm) | $\Sigma$ (mm) | $\sigma$ (mm) |
| --- | --- | --- | --- |
| x (left/right) | -0.3 | 3.3 | 1.7 |
| y (sup/inf) | 0.7 | 2.6 | 2.9 |
| z (ant/post) | 0.1 | 1.4 | 2.0 |

### 3.3 Comparison of Reference Plan and Daily Adaptive Plans

The summed dose to the clinical target volume (CTV) was within [-0.61%, 1.78%] of the reference plan dose in all cases, with the summed dose falling below the reference plan dose in only 2/10 cases (Figure 2). For the contralateral carotid artery, ipsilateral parotid gland, contralateral parotid gland, contralateral cochlea, and brainstem, the summation plan dose was lower than the reference plan dose in all cases. The dose to the ipsilateral cochlea was higher in the summation plan for 3/7 cases, but all cases fell within a maximum dose deviation of 4.3%. Doses to the spinal cord exceeded those in the reference plans for cases 1 and 2 by 8.1% and 2.1%, respectively. However, the planning constraint for the spinal cord used in these two cases was D_max_ < 10 Gy rather than the 45 Gy dose constraint used in cases 3–10. The total summed doses were between 10 and 11 Gy in these two cases, and these small absolute deviations in dose resulted in higher percent deviations despite the doses falling far below the typical constraint of 45 Gy. Similarly, dose sparing of 5%–12% was seen for five OARs in case 4, which was the largest dose sparing benefit seen for any case. Case 4 only had four fractions delivered, so the percent differences are larger than measured in most other cases.

**Figure 2:**
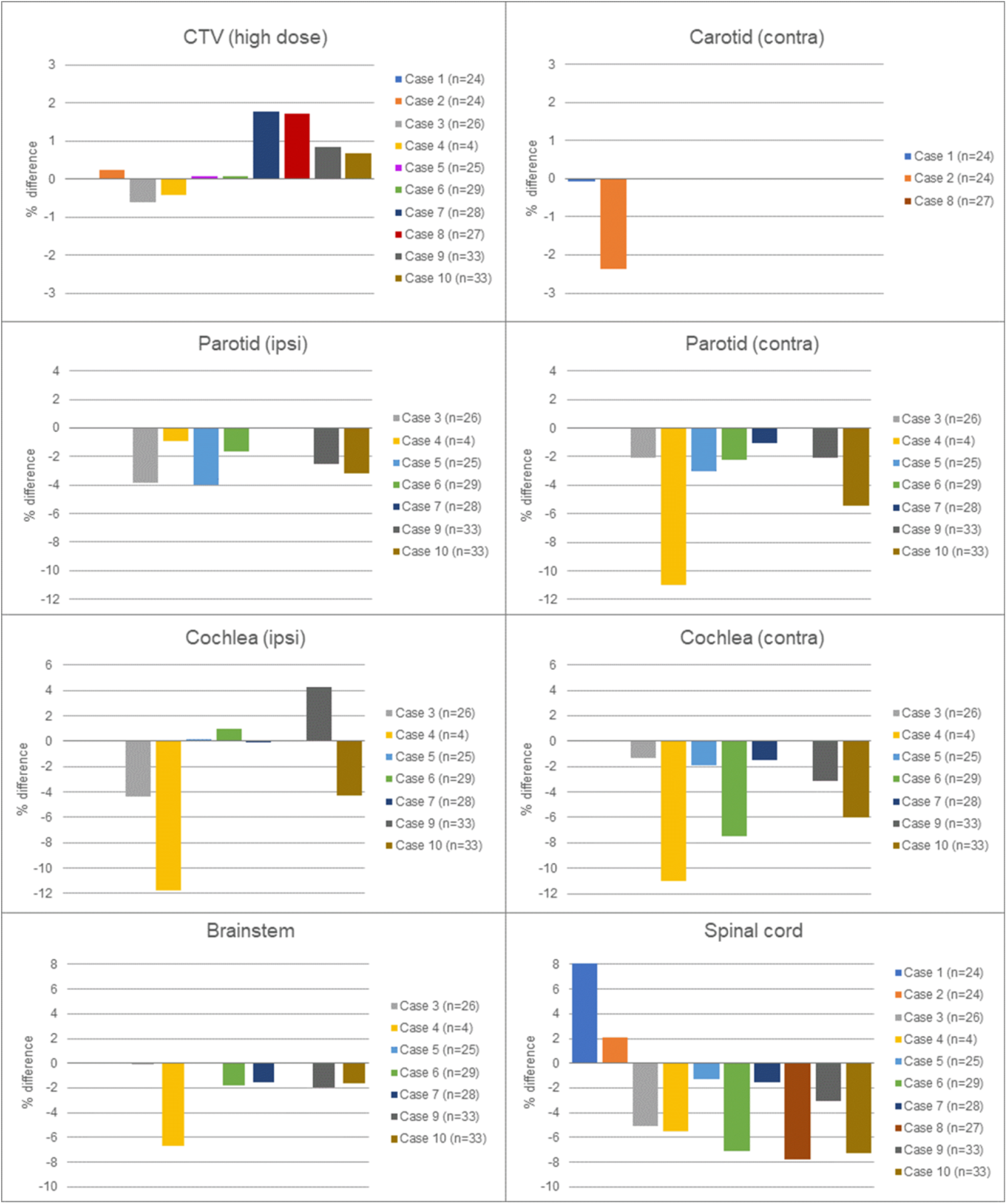
Percent differences between the summation plan and reference plan doses for each plan quality metric specified in Appendix D. Positive values mean that the dose in the summation plan exceeded the dose in the reference plan. The number of treatments delivered on the MR-linac (n) for each case is provided in the legends. In cases where no dosimetric criterion was used for an OAR, that case was omitted from the legend of the graph. “Ipsilateral” and “contralateral” are abbreviated “ipsi” and “contra,” respectively.

All cases had at least two fractions where one or more planning constraint was not met in the adaptive plan (Table 3). However, in most cases where a constraint was violated in at least one fraction, that constraint was still met in the summation plan. For the ipsilateral parotid glands in cases 3, 6, and 9 and the contralateral parotid gland in case 3, the constraints were not met in the initial reference plan and therefore were violated at nearly every fraction and also in the summation plan. There were only two instances (spinal cord in case 1 and ipsilateral cochlea in case 9) where the constraint was met in the reference plan but not in the summation plan. As previously mentioned, the constraint used for the spinal cord in case 1 was D_max_ < 10 Gy rather than the typical 45 Gy, so this constraint was not heavily enforced during on-line plan adaptation.

**Table 3:**
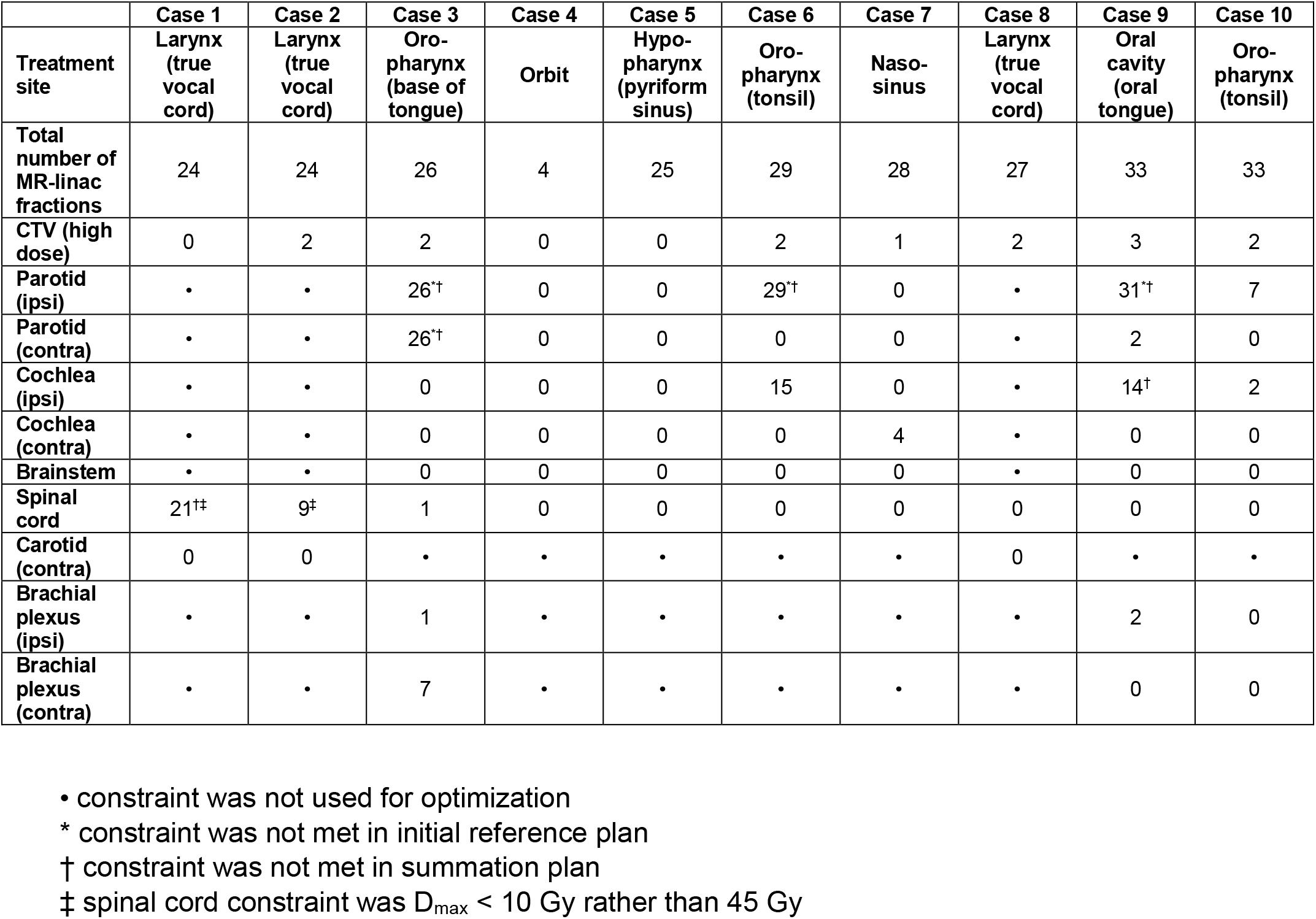
Number of times a constraint was violated in an adaptive plan.

Case 9 was chosen as a representative case to illustrate the dosimetric differences between the expected cumulative dose (if the reference plan had been delivered daily) and the actual cumulative dose delivered to the OARs and CTV (Figure 3). Despite the fluctuations in dose between adaptive treatments, minimal deviation was observed between the delivered and expected doses from fraction to fraction (within [-122.7 cGy, 152.4 cGy] over all cumulative fractions). The largest deviation occurred for the ipsilateral cochlea, which had a total of 14 constraint violations out of 33 fractions, resulting in a total dose deviation of 141.8 cGy (4.3%) at the end of treatment. Overall, doses to all OARs were highly consistent between the reference plan and summation plan in all cases, despite constraint violations and variations in daily dose distributions due to on-line plan adaptation.

**Figure 3:**
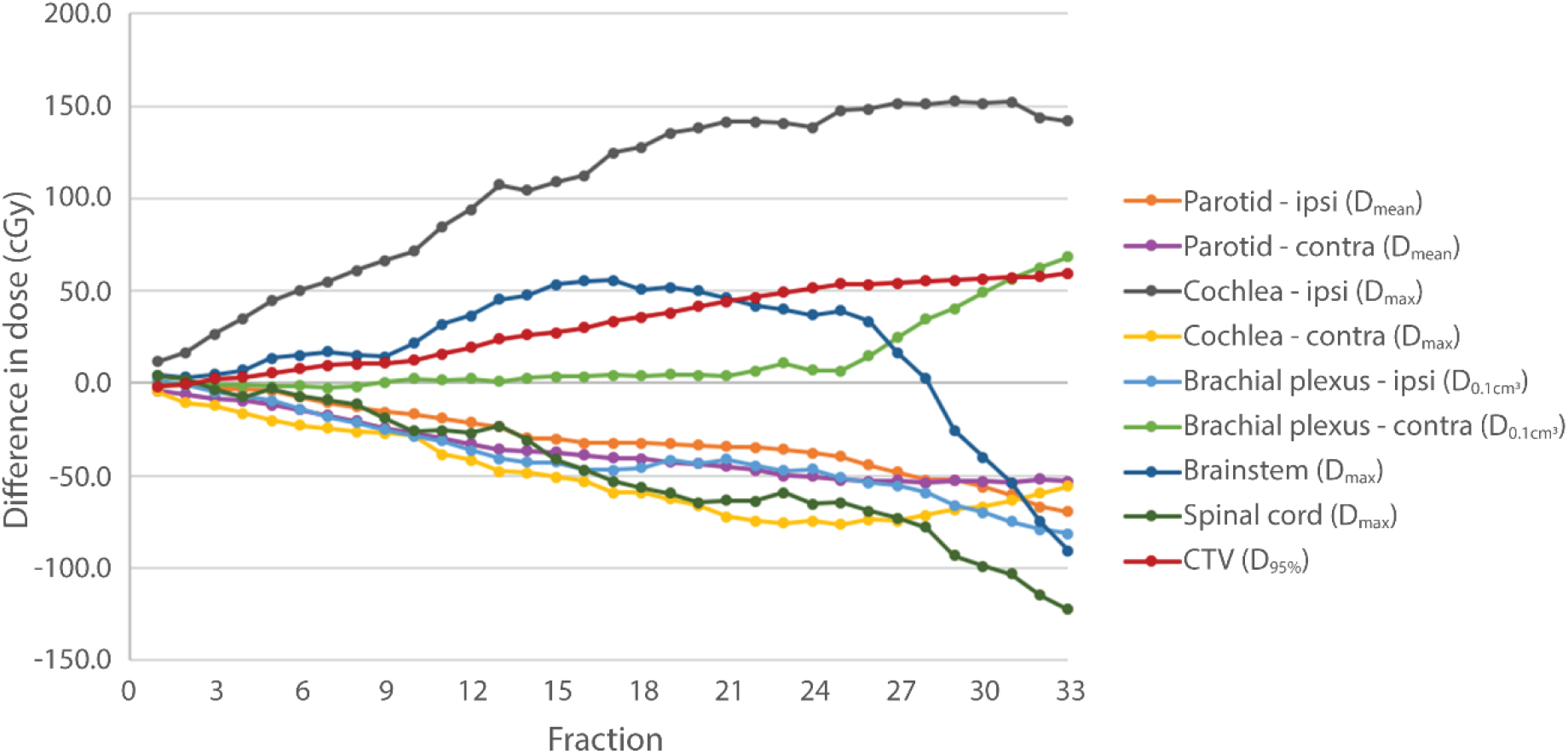
The absolute difference between the cumulative delivered dose and the expected delivered dose if the reference plan had been delivered at each fraction for case 9 (treatment site: oral tongue). Positive values mean that the cumulative delivered dose exceeded the reference plan expected dose. An off-line ATS was performed between fractions 25 and 26. This case was chosen as a representative case because all fractions were delivered on the MR-linac.

## 4. Discussion

ART has begun to play an increasingly prominent role in the treatment of HNC for two primary reasons: 1) significant tumor shrinkage, weight loss, and large anatomical deformations of OARs are often observed for HNC patients during RT [16–18]; and 2) treatment-related side effects from conventional RT can be particularly debilitating for HNC and often persist for years after treatment [19,20]. Several studies have demonstrated clinically significant reductions in doses to the parotid glands [21–23], spinal cord [23,24], and swallowing-related structures [25,26] with one or more off-line plan adaptations over the course of radiotherapy. Because off-line adaptive re-planning is time- and resource-intensive, many studies have focused on identifying anatomical and dosimetric guidelines to determine when plan adaptation would be optimally effective [24,25,27]. However, the clinical introduction of the 1.5T MR-linac has made daily ART for HNC a feasible clinical reality.

In this study, we present our initial experience with daily MR-guided ART for HNC on a 1.5T MR-linac. We employed an ART_ex_aequo_ approach [14] intended to deliver prescription and OAR doses defined prior to therapy. Target volumes were not modified, and OAR contours were updated only when a new reference plan was created via an off-line ATS plan adaptation. We used this conservative approach for our first HNC case series on the MR-linac to test safety and feasibility and to iteratively improve our clinical workflow. However, following the present R-IDEAL stage 2a/2b systematic evaluation, we plan to transition to ART approaches intended to spare OAR doses and/or handle shrinking tumor volumes in the near future [28]. The technological innovations with the MR-linac platform, including enhanced visualization of soft tissue and the on-line ATP and ATS workflows, have the potential to overcome many of the time and resource limitations associated with OAR-sparing and target volume-modifying ART approaches.

Total treatment times ranged from 31-85 minutes, with 91% completed within 60 minutes. Most of the upper outliers were due to isocenter shifts being too large or the on-line adaptive plan not meeting acceptance criteria. In these cases, the patient was repositioned and may have been given a short break out of the mask. Although these treatment durations are longer than HNC treatments on conventional linacs, they are comparable to other specialized image-guided procedures in our clinic.

One common concern with on-line adaptive re-planning is that a plan must be delivered before patient-specific IMRT QA is performed, in significant contrast to conventional QA workflows. However, all adaptive treatment plans in this study passed IMRT QA (γ>90%), with 75% of plans scoring 99.4% or higher. These high pass rates may be attributed to the comprehensive series of safety checks that are implemented at various stages of the clinical workflow, including annual, monthly, and daily linac and MRI QA as well as the secondary MU check during on-line plan adaptation. This workflow enables any dosimetric issues to be caught long before the patient is treated and ensures that on-line adaptive treatments are safe for all patients.

The setup variability results indicate that immobilization in a custom thermoplastic head and neck mask resulted in highly reproducible patient positioning with setup errors consistent with values previously reported in the literature [29–31]. In both the ATP and ATS workflows of the Unity system, an adaptive plan cannot be created if the isocenter shift exceeds 5 cm in any one direction because the table can only be moved longitudinally along the y (superior/inferior) axis. All isocenter shifts fell far below this threshold, with maximum shifts of 0.80, 1.97, and 0.73 cm in the x, y, and z directions over all 253 adaptive plans. Although shifts greater than 1 cm seem quite large with a mask, this occurred only four times and only in the y direction, which can be attributed to errors in positioning the mask along the index bar. In these cases, a decision was made not to reposition the patient because an acceptable ATP plan could still be generated. However, while on-line plan reoptimization directly accounts for inter-fraction positional variations, minimizing these variations can help the optimization algorithms achieve “better plans” that meet all dosimetric constraints.

Differences between the reference plan doses and summed doses were minimal, with the largest percent deviations occurring when the absolute doses were small (as in the case of the spinal cord D_max_ < 10 Gy constraint or when only 4 fractions were delivered). It is important to note that the ATP workflow was used for the majority of fractions, with only a single ATS fraction for cases 6, 8, 9 and 10. There was no modification of the CTV or intentional dosimetric sparing to any OARs for these cases. Any dosimetric sparing to the OARs is likely caused by small changes in the locations of “hot spots” in the dose distribution within each OAR, which is a result of both daily plan adaptation and the inherent randomness of the Monte Carlo dose calculation. Instead of the “hot spot” being located in the same voxel at each fraction, the “hot spot” is smeared around a larger volume over all fractions, effectively reducing the maximum or mean cumulative dose to any OAR compared to the reference plan. Overall, the consistency between the reference and summation plan doses demonstrate that our conservative initial clinical implementation of the ATP workflow on the MR-linac for HNC is safe for patients and performs at least as well as the standard of care from a dosimetric perspective.

One drawback of this dosimetric analysis was that the adaptive plans were summed using rigid plan summation on the anatomy of the reference plan in the ATP workflow. This dose summation approach was necessary because there are currently no validated deformable dose accumulation tools for the Unity system. In the absence of such a tool and with only ten cases in this initial feasibility study, it is difficult to draw conclusions about the dose sparing benefit of this MR-guided ART_ex_aequo_ approach. However, the current standard of care in RT is to plan multi-fraction treatments based on the obviously erroneous assumption that the anatomy remains constant throughout all treatment fractions. Thus, all dose estimates in conventional RT are based on the simulation image, which is analogous to the reference plan anatomy in MR-guided daily adaptive RT. Because all doses were summed on the reference plan anatomy, we may conservatively conclude that the dosimetric variability between daily adaptive plans results in minimal variation from the reference plan over the course of RT.

Despite these limitations, this effort represents the first report of 1.5T MR-guided daily ART for HNC and the first prospective series of dosimetric analyses of HNC treatment on a hybrid MR-linac system. Consistent with R-IDEAL, in this planned “prospective small uninterrupted case series,” we have demonstrated “technical improvements, feasibility, and safety” [11] of 1.5T daily ART_ex_aequo_ treatment for HNC. The continued R-IDEAL programmatic integration of this data with extant efforts across the MR-Linac Consortium is designed to ensure that rather than *ad hoc* reportage, iterative technical developments on a new technology are reported systematically. Future prospective analyses combining HNC data from other MR-Linac Consortium sites using the same workflow are currently being planned. Furthermore, first technical studies have shown that MR imaging using the 1.5 T Unity system not only provides excellent anatomical image data [32] but also allows for robust and reproducible quantitative imaging techniques, such as DWI [33]. Consequently, functional MR data acquired sequentially during MR-guided RT needs to be investigated systematically in order to pave the way towards future biologically individualized RT of HNC. Next steps include an R-IDEAL Stage 2a evaluation of functional imaging integration and feasibility with DWI, and Stage 2a implementation using an ART_reduco_ [14] approach within the prospective Phase II randomized MR-ADAPTOR trial [28].

## 5. Conclusion

The aim of this article was to describe our institution’s clinical workflow for treating HNC on a 1.5T MR-linac and to demonstrate safety and feasibility through a prospective analysis of ten cases treated with an on-line ATP workflow. Treatment times remained under one hour in 91% of cases. All treatment plans passed IMRT QA, and patient immobilization in a custom head and neck mask resulted in highly reproducible patient setups. The daily adaptive plans were very consistent with the reference plans with minimal dosimetric differences. In summary, daily MR-guided adaptive RT for HNC is safe and clinically feasible with the MR-linac, although further studies with more advanced dose accumulation strategies are required to investigate the true dosimetric impact of this treatment approach.

## Data Availability

The data used for this analysis will be posted on NIH Figshare in the coming weeks. A data descriptor publication is currently being prepared.

## Appendix A

*Head and Neck Clinical and Technical Profile*

This document was developed as a multi-institutional collaborative effort by the MR-Linac Consortium Head and Neck Tumor Site Group. The information below represents the iteration as of June 2020.

### Clinical and Technical Profile Definitive Oropharyngeal Squamous Carcinomas, with or without Concurrent Chemotherapy (non-adaptive/standard of care)

#### 1. Abstract

The proposed template is included for **standard of care** (i.e. doses of 200-212 cGy) delivered to a non-shrinking planning target volume (PTV), for head and neck squamous carcinomas of the oropharynx. Oropharyngeal cancer is defined as two distinct etiologic entities: human-papilloma virus positive (HPV+) squamous carcinomas, which have a notably more radiosensitive biological response, with expected 5-year survival of 85% or greater in aggregate, and human-papilloma virus negative (HPV-) carcinomas, which portend substantially worse survival. The publication for low-intermediate risk (AJCC 7^th^ edition) of the DE-ESCALATE [1] trial demonstrated 2-year overall mortality of 2.5% and 2-year locoregional recurrence rate of 6%. Similarly, RTOG 1016 [2] showed a 5-year survival of 84.6% with concurrent 3-week cisplatin. Consequently, this profile is designed for the implementation of similar dosing schema, with concurrent cisplatin, for both HPV+ and HPV-head and neck squamous carcinomas. However, based on institutional policy, wherein published data have shown for exceedingly low-risk cases, a <5% 5-year probability of any event (mortality or recurrence/metastasis) for selected AJCC 7^th^ edition T1-2N0-2bM0/AJCC 8^th^ edition T1-2N1M0 cases, where chemotherapy is omitted, this profile shall note operational variance.

For extended development of Consortium-supported oropharynx clinical trial, please see Bahig et al. [3] for full clinical therapy, planning, OAR, segmentation, and treatment details.

#### 2. Describe the clinical cohort of interest

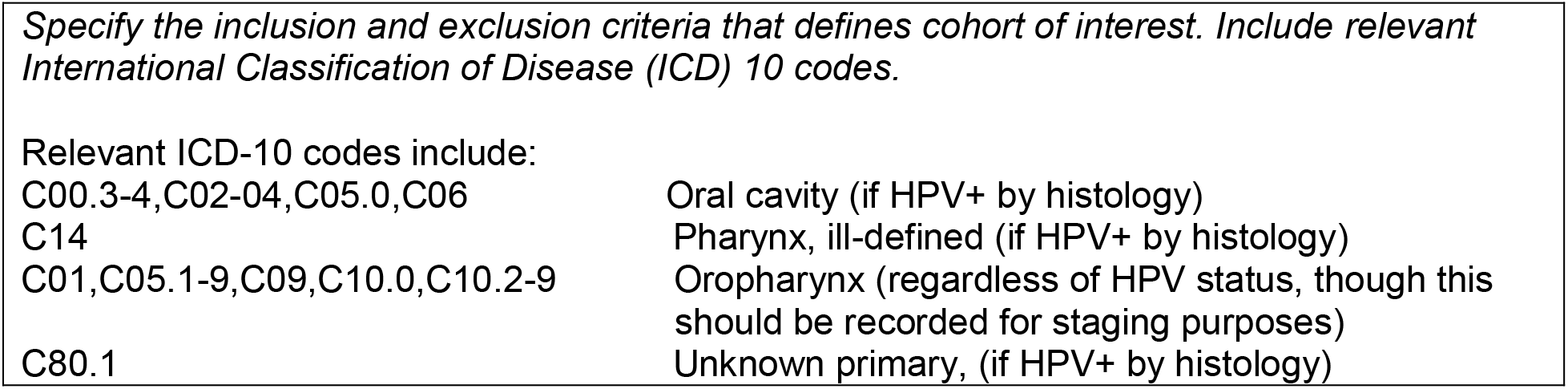

#### 3. Describe any pre-simulation patient preparation

-Patients are recommended to have pre-simulation formal dental examination, evaluation, and extraction/renovation with interval healing before simulation.

-Patients are recommended to have consideration for construction of an integrated apparatus for immobilization/fixation of the dentition for purposes of immobilization, as well as context specific dental apparatus to displace or shield normal tissue. Examples include, but are not limited to an integrated bite-block, manually constructed “tongue-blade” or “cork and blade” devices, or more elaborate custom dental stents. Lateralizing, medializing, tongue depressing, or maxillary/palatal fixation should be carefully considered on an individual case-basis, with an eye towards dose minimization.

#### 4. Describe patient simulation

4. Describe patient simulation.
|  |  |
| --- | --- |
| Body Positioning: | Supine |
| Head positioning: | Neural |
| Arm positioning: | By side, on chest, or folded |
| Other positioning: |  |
| Patient Positioning Devices: | Headrest (acceptable); Occipital immobilization cradle (preferred) |
| Bolus: | None |
| CT (PET/CT, other) extent | Vertex to carina (standard); PET-CT simulation in mask is preferred if staging PET has not been performed prior. If a separate staging PET-CT (non-mask) is performed, deformable image-registration is encouraged. CT in the same mask position is required for dose calculation, and should use comparable slice thickness reconstruction to the MRL-simulation scan. |
| CT (PET/CT, other) slice thickness (mm) | $\leq 2.5$ |
| Contrast | IV contrast (preferred) |
| 4D extent and goals of assessment | None |
| Isocenter location | Within the target volume; at hyoid. |
| Other: | -For ATL2/Unity current version, a maximum superior-inferior treatment field length of 22 cm is extant, necessitating a maximum acceptable S/I distance on $\leq 21$ cm ($\leq 20$ cm preferred) after PTV expansion.<br>-Ideally, MR- and CT-simulation should occur in $<24$ hours; $>7$ day intervals between CT- and MR-simulation scans should be eschewed.<br>-Contrast utilization (CT or MRI) is encouraged; MRI contrast is preferred.<br>-In-mask simulation CT, PET, and MR imaging performed in less than 7-day interval should be rigidly co-registered.<br>-EPI-based diffusion-weighted imaging (DWI) may be c-registered, and may be used qualitatively for reference, but should not be registered for voxel-specific segmentation within the standard of care profile. |

#### 5. Describe Simulation MRI sequence(s)

5. Describe Simulation MRI sequence(s) MR-Linac sequences:
|  | T2w - 2 min | T1w - 2 min | T2w - 6 min | T1w - 6 min | DWI (b=0, 150, 500) | ADC map |
| --- | --- | --- | --- | --- | --- | --- |
| MR manufacturer | Philips | Philips | Philips | Philips | Philips | Philips |
| Field Strength | 1.5 T | 1.5 T | 1.5 T | 1.5 T | 1.5 T | 1.5 T |
| Sequence name | T2 3D Tra 2min | T1 3D Tra 2min | T2 3D Tra | T1 3D Tra | DWI_3b-SPAIR FO RL | dADC_3b-SPAIR SENSE |
| Coil | Multi coil | Multi coil | Multi coil | Multi coil | Multi coil |  |
| Slice thickness (mm) | 2 | 2 | 2.2 | 2.4 | 4 | 4 |
| Resolution x/y plane (recon.) (mm) | 0.83 | 0.83 | 0.68 | 0.68 | 1.25 | 1.25 |
| Slice gap (mm) | 1 | 1 | 1.1 | 1.2 | 4 | 4 |
| Bandwidth (Hz/pixel) | 740 | 432 | 459 | 344 | 1930 |  |
| FOV (mm) | 400 | 400 | 520 | 380 | 400 | 400 |
| No. of slices | 300 | 300 | 264 | 208 | 75 (x3) | 75 |
| Time to echo (ms) | 277.818 | 4.604 | 375 | 4.501 | 68.09 |  |
| Time to repetition (ms) | 1535 | 11 | 2100 | 13 | 14432.76 |  |
| Flip angle (degrees) | 90 | 30 | 90 | 27 | 90 |  |
| Slice acquisition | 3D | 3D | 3D | 3D | 2D |  |
| Sequence type | SE | GR | SE | GR | SE |  |

Optional With/Without Contrast Pre-/Mid/Post-therapy Simulation Sequences (part 1):
|  | T2w | T1w | T1 map (2, 5, 10, 15, 20°) | DCE (x56) | Dixon_op p | Dixon_in | Dixon_F | Dixon_W |
| --- | --- | --- | --- | --- | --- | --- | --- | --- |
| MR manufacturer | Siemens | Siemens | Siemens | Siemens | Siemens | Siemens | Siemens | Siemens |
| Field Strength | 1.5 | 1.5 | 1.5 | 1.5 | 1.5 | 1.5 | 1.5 | 1.5 |
| Sequence name | T2 FS TSE (For TPS) | Ax 3D T1 (For TPS) | t1_map_n deg | +C DCE_56dyn | +C Ax 3D T1 Dixon (For TPS)_opp | +C Ax 3D T1 Dixon (For TPS)_in | +C Ax 3D T1 Dixon (For TPS)_F | +C Ax 3D T1 Dixon (For TPS)_W |
| Coil | Posterior/bilateral flex | Posterior/bilateral flex | Posterior/bilateral flex | Posterior/bilateral flex | Posterior/bilateral flex | Posterior/bilateral flex | Posterior/bilateral flex | Posterior/bilateral flex |
| Slice thickness (mm) | 2 | 1 | 4 | 4 | 1 | 1 | 1 | 1 |
| Resolution x/y plane (recon.) (mm) | 0.5 | 0.5 | 1 | 1 | 1 | 1 | 1 | 1 |
| Bandwidth (Hz/pixel) | 300 | 400 | 385 | 500 | 405 | 405 | 405 | 405 |
| FOV (mm) | 256 | 256 | 256x208 | 256x208 | 256 | 256 | 256 | 256 |
| No. of slices | 120 | 240 | 30 (x5) | 30 (x56) | 240 | 240 | 240 | 240 |
| Time to echo (ms) | 80 | 4.77 | 1.27 | 1.09 | 2.39 | 4.77 | 2.39 | 2.39 |
| Time to repetition (ms) | 5280 | 7.38 | 5 | 5 | 7.11 | 7.11 | 7.11 | 7.11 |
| Flip angle (degrees) | 180 | 19.0765 | 2, 5, 10, 15, 20 | 17.3417 | 10 | 10 | 10 | 10 |
| Slice acquisition | 2D | 3D | 3D | 3D | 3D | 3D | 3D | 3D |
| Sequence type | SE | GR | GR | GR | GR | GR | GR | GR |

Optional With/Without Contrast Pre-/Mid/Post-therapy Simulation Sequences (part 2):
|  | DWI Blade (b=0, 800) | ADC map | T2 map (14 echoes) | DWI Resolve (b=0, 200, 400, 600, 800, 1000) | ADC map |
| --- | --- | --- | --- | --- | --- |
| MR manufacturer | Siemens | Siemens | Siemens | Siemens | Siemens |
| Field Strength | 1.5 | 1.5 | 1.5 | 1.5 | 1.5 |
| Sequence name | Ax DWI Blade b0_800_TRACE W | Ax DWI Blade b0_800_ADC | T2 mapping 14 echoes_T2Map | DWI resolve_ivim_sms_TRACEW | DWI resolve_ivim_sms_ADC |
| Coil | Posterior/bilateral flex | Posterior/bilateral flex | Posterior/bilateral flex | Posterior/bilateral flex | Posterior/bilateral flex |
| Slice thickness (mm) | 4 | 4 | 4 | 4 | 4 |
| Resolution x/y plane (recon.) (mm) | 2 | 2 | 1 | 1 | 1 |
| Slice gap (mm) | 4 | 4 | 4 | 4 | 4 |
| Bandwidth (Hz/pixel) | 1220 |  | 250 | 750 |  |
| FOV (mm) | 256 | 256 | 256 | 256 | 256 |
| No. of slices | 27 (x2) | 27 | 48 | 24 (x7) | 24 |
| Time to echo (ms) | 50 |  | 10 | 68 |  |
| Time to repetition (ms) | 5900 |  | 4240 | 1500 |  |
| Flip angle (degrees) | 120 |  | 180 | 180 |  |
| Slice acquisition | 2D |  | 2D | 2D |  |
| Sequence type | SE |  | SE | RM |  |

#### 6. Describe Organs at Risk (OAR), Gross Tumor Volumes (GTVs), Clinical Target Volumes (CTVs), Internal Target Volumes (ITVs), Planning Organ at Risk Volume (PVR), and Planning Target volumes (PTVs)

OAR definitions should be consistent with AAPM Task Group 263 nomenclature and identify the Foundational Model of Anatomy (FMAID) number when available. The following guidelines are used for specifying superior, anterior, posterior, right lateral, left lateral and inferior borders of OARs and CTVs:

‐ OARs: Brouwer et al. [4]
‐ Primary tumor CTVs: Gregoire et al. (2018) [5]
‐ Neck node levels: Gregoire et al. (2014) [6]

For all target volumes, omnidirectional CTV-to-PTV margins should be constructed via the Van Herk [7] formula of: 2.5∑ + 0.7σ. For OAR-to-PRV margins, the McKenzie [8] formula of = 1.3∑ + 0.5σ may be used. Variable margin strategies such as Yang et al. [9] are encouraged for patients with head and neck cancer.

#### 7. Contouring Atlases for off-line auto-segmentation

The following contouring atlases are consistent with CTP contouring guidelines described above that can be used for off-line auto-segmentation. These are stored in an online repository.

See the following consensus guidelines (weblink):

‐ https://www.eortc.org/rog-guidelines/
‐ https://www.rtog.org/CoreLab/ContouringAtlases/HNAtlases.aspx

#### 8. Describe radiation prescription

| Phase | Cumulative |
| --- | --- |
| PTV (High Risk) | PTV_6996; PTV_7000 |
| PTV (High Risk) Dose per fraction | 200-212 cGy |
| PTV (Intermediate Risk) [optional] | PTV_6600; PTV_6300 |
| PTV (Intermediate Risk) Dose per fraction [optional] | 180-200 cGy |
| PTV (Low Risk) | PTV_4000; PTV_5000,PTV_5600 |
| PTV (Low Risk) Dose per fraction | 180-200 cGy |
| Number of fractions | 33/35 |
| Fractions per day | 1 (5 or 6 fractions weekly) |
| Total dose in phase | 6996/7000 cGy |
| Cumulative dose with prior phases | 6996/7000 cGy |
| Delivery technique | Step and shoot IMRT(acceptable), VMAT (preferred) |

#### 9. Describe initial treatment planning

Full planning constraints used for oropharynx patients on the MR-ADAPTOR protocol are detailed as per included trial documentation.

For non-trial cases, the following goals are used for specified OARs:

‐ Spinal cord-Max 45 Gy
‐ Spinal cord PRV (5mm)-Max 48 Gy
‐ Brainstem-V30<30%; Max <40Gy
‐ Brain-<1cm^3^ receiving 54 Gy
‐ Mandible-V40 <40%; Max dose less than 70 Gy
‐ Contralateral parotid-Mean <26 Gy; V15<50%.

Contralateral Submandibular gland (if uninvolved)-Mean <26 Gy; V15<50%.

Ipsilateral Submandibular gland (if uninvolved)-Mean <39 Gy; V25<50%.

‐ Brachial plexus-Max <66 Gy
‐ Esophagus-Mean <30 Gy
‐ Larynx-Mean <30 Gy

Oral cavity-Mean <30Gy

#### 10. Identify pre-beam set up, beam-on and post-beam MR Sequence(s)

Patients are immobilized in a occipital cradle (Klarity) with S-frame thermoplastic mask and integrated bite-block/stent as per Wang et al. [10] and Ding et al. [11].

Pre-therapy CT and MR simulation is performed on conventional scanners, with 2-3mm slice thickness reconstructions. An MRL-specific simulation is performed on the MRL using the Elekta/Philips-provided T1, T2, and EPI DWI full-length sequences, as well as the “short” (2-minute) T2 sequence for daily alignment. Rigid CT-MR fusion is performed as per Kiser et al. [12].

A 2-minute T2 scan is used for alignment. After Monaco/Unity registration, an action level is applied as follows:

‐ All shifts in all axes <3mm: Proceed to ATP workflow.
‐ Any axis >3 but <5mm: Physician on-line review of images; if approved, proceed to ATP workflow.
‐ Any axis >5mm: Reposition patient; if still >5mm, MD call to table for process review.

If shift is >5mm, consider ATS online, or offline ATS with next day ATP. ATS is preferred offline, with daily ATP, 2/2 current time-on-table constraints for OPC cases.

‐ If all pre-therapy OAR/TV goals are met, therapy can proceed.
‐ If any goals are not met, real-time MD review is required before treatment.

Beam-on sequence currently uses 2D orthogonal motion management package, but desperately needs DICOM export and quantification to be useful.

Post-beam EPI DWI is currently collected; we will be optimizing sequences through research protocols.

#### 11. Describe daily registration

Daily registration utilizes hyoid as a surrogate structure, with spatial cord, mandible, GTV and parotids used to evaluate post-action-level alignment.

#### 12. Describe daily plan adaptation strategy

Current default strategy is position compensation owing to time constraints. Initial plan adaptation strategy uses ATS and segment weight optimization. If all pre-therapy constraints are met, treatment can proceed; otherwise, real-time online MD approval is required. Weekly dose summation is used for physics review; if all pre-therapy constraints are met, treatment can proceed; if a constraint is exceeded, offline ATS is performed, approved by MD is performed, and used as the base for ATP the following day.

#### 13. Describe beam-on motion management strategy, if applicable

None to date

#### 14. Describe commonly encountered delivery issues and mitigation strategies

‐ We routinely have found for head and neck cases that ATS replanning can take >10 minutes, and the cumulative time in mask can be uncomfortable for patients; consequently, we routinely plan ATS offline, and use ATP on the offline ATS approach the next day.
‐ We are unable to output motion monitoring images form the proprietary format to DICOM; we consequently cannot perform action (online or offline) other than visual inspection of swallowing motion.

## Appendix B

### MR-Linac Patient Evaluation Form

Patient MRN:___________

Tx Site: ___________

Requested total dose (cGy): ___________

No of Fractions: ___________

Requesting Physician: ___________

Evaluating Physician: ___________

Evaluating Physicist: ___________

### PATIENT CONSIDERATIONS

1. Does patient have any MR contraindications - metal objects (implanted/on person), pacemakers, defibrillators, neurostimulators …? Yes No
2. Max patient height (lying down on table) from table top (cm): _______ (Max = 36.5)
3. Max patient width (lying down on table) from table top (cm): ______ (Max = 63.5)
4. Is patient self-ambulatory? Yes No
5. Is patient claustrophobic? Yes No
6. Can patient hold arms raised above shoulders, if needed (∼60 mins)?: Yes No
7. Can patient hold bladder for approx. 1 hour? Yes No
8. Does the target extent (PTV) exceed 20 cm along ‘long’ direction? Yes No
9. Does the target extend laterally to > 20 cm from midline? Yes No
10. Can physician/physics provide coverage for all fractions during Tx? Yes No
11. Does no of treatment isocenters exceed 1? Yes No
12. Can the patient/target be immobilized using immobilization aids? Yes No

### SPECIAL CONSIDERATIONS

1. Motion management (Currently, cannot automatically turn ON/OFF beam)
  a. FB ITV based approach
  b. Breath Hold
  c. 4D CT?
2. Contrast MR? For sim only or daily?
3. Other Special immobilization aids?

## Appendix C

### MRI Patient Safety Screening Form

1. Do you have a pacemaker or defibrillator?
2. Do you have a spinal cord stimulator?
3. Do you have an implanted infusion pump?
4. Do you have ear implants?
5. Do you have retinal implants?
6. Do you have a Port/Port-a-Cath?
7. Can you lie flat and remain still?
8. Does being in an enclosed space make you nervous or fearful?
9. Who provided this information (if not the patient?)

*This screening form is filled out every time the patient arrives at the MR-Linac.

## Appendix D

**Table D.1:** MRI protocol parameters for the standard Unity T2-weighted sequences. The 6-minute scan is from the Unity Plan for Care for Head and Neck, and the 2-minute scan is from the Unity Plan of Care for Pelvis.

|  | **2-minute T2** | **6-minute T2** |
| --- | --- | --- |
| Field of view (mm): RL x AP x FH | 400 x 400 x 300 | 520 x 520 x 250 |
| Acquisition voxel size (mm): RL x AP x FH | 1.5 x 1.5 x 2.0 | 1.2 x 1.2 x 2.2 |
| Reconstruction voxel size (mm): RL x AP x FH | 0.83 x 0.83 x 1.0 | 0.68 x 0.68 x 1.1 |
| Slice acquisition mode | 3D | 3D |
| Scan technique | Turbo spin echo | Turbo spin echo |
| Echo train length | 114 | 150 |
| Echo time (ms) | 278 | 375 |
| Repetition time (ms) | 1535 | 2100 |
| Flip angle (deg) | 90 | 90 |
| Refocusing control angle (deg) | 100 | 100 |
| Bandwidth (Hz/pixel) | 740 | 459 |
| Scan duration (min:sec) | 1:57 | 6:03 |

## Appendix E

**Table E.1:** Dosimetric constraints/plan quality metrics used for all patient plans. All numerical values are in cGy unless otherwise specified. “Ipsilateral” and “contralateral” are abbreviated “ipsi” and “contra,” respectively. A • symbol means that no dosimetric constraint was used for that OAR.

|  | Case 1 | Case 2 | Case 3 | Case 4 | Case 5 | Case 6 | Case 7 | Case 8 | Case 9 | Case 10 |
| --- | --- | --- | --- | --- | --- | --- | --- | --- | --- | --- |
| Treatment site | Larynx (true vocal cord) | Larynx (true vocal cord) | Oropharynx (base of tongue) | Orbit | Hypopharynx (pyriform sinus) | Oropharynx (tonsil) | Nasosinus | Larynx (true vocal cord) | Oral cavity (oral tongue) | Oropharynx (tonsil) |
| CTV (high dose) | $D_{90\%} > 6300$ | $D_{90\%} > 6300$ | $D_{95\%} > 6996$ | $V_{1400\text{cGy}} > 98\%$ | $D_{95\%} > 6996$ | $D_{95\%} > 6996$ | $V_{6000\text{cGy}} > 90\%$ | $D_{90\%} > 6525$ | $D_{95\%} > 6996$ | $D_{95\%} > 6996$ |
| Parotid (ipsi) | • | • | $D_{\text{mean}} < 2600$ | $D_{\text{mean}} < 2600$ | $D_{\text{mean}} < 2600$ | $D_{\text{mean}} < 2600$ | $D_{\text{mean}} < 2600$ | • | $D_{\text{mean}} < 2600$ | $D_{\text{mean}} < 2600$ |
| Parotid (contra) | • | • | $D_{\text{mean}} < 2600$ | $D_{\text{mean}} < 2600$ | $D_{\text{mean}} < 2600$ | $D_{\text{mean}} < 2600$ | $D_{\text{mean}} < 2600$ | • | $D_{\text{mean}} < 2600$ | $D_{\text{mean}} < 2600$ |
| Cochlea (ipsi) | • | • | $D_{\text{max}} < 3500$ | $D_{\text{max}} < 3500$ | $D_{\text{max}} < 3500$ | $D_{\text{max}} < 3500$ | $D_{\text{max}} < 3500$ | • | $D_{\text{max}} < 3500$ | $D_{\text{max}} < 3500$ |
| Cochlea (contra) | • | • | $D_{\text{max}} < 3500$ | $D_{\text{max}} < 3500$ | $D_{\text{max}} < 3500$ | $D_{\text{max}} < 3500$ | $D_{\text{max}} < 3500$ | • | $D_{\text{max}} < 3500$ | $D_{\text{max}} < 3500$ |
| Brainstem | • | • | $D_{\text{max}} < 5400$ | $D_{\text{max}} < 5400$ | $D_{\text{max}} < 5400$ | $D_{\text{max}} < 5400$ | $D_{\text{max}} < 5400$ | • | $D_{\text{max}} < 5400$ | $D_{\text{max}} < 5400$ |
| Spinal cord | $D_{\text{max}} < 1000$ | $D_{\text{max}} < 1000$ | $D_{\text{max}} < 4500$ | $D_{\text{max}} < 4500$ | $D_{\text{max}} < 4500$ | $D_{\text{max}} < 4500$ | $D_{\text{max}} < 4500$ | $D_{\text{max}} < 4500$ | $D_{\text{max}} < 4500$ | $D_{\text{max}} < 4500$ |
| Carotid (contra) | $D_{\text{mean}} < 1000$ | $D_{\text{mean}} < 1000$ | • | • | • | • | • | $V_{3500\text{cGy}} < 5\%$ | • | • |
| Brachial plexus (ipsi) | • | • | $V_{6000\text{cGy}} < 1 \text{ cm}^3$ | • | • | • | • | • | $D_{0.1\text{cm}^3} < 6600$ | $D_{\text{max}} < 6600$ |
| Brachial plexus (contra) | • | • | $V_{6000\text{cGy}} < 1 \text{ cm}^3$ | • | • | • | • | • | $D_{0.1\text{cm}^3} < 6600$ | $D_{\text{max}} < 6600$ |
| Brain | • | • | • | • | • | • | • | • | • | $D_{\text{max}} < 5400$ |
| Mandible | • | • | • | • | • | • | • | $D_{\text{max}} < 6500$ | • | • |
| Lens (ipsi) | • | • | • | • | • | • | $D_{\text{max}} < 1000$ | • | • | • |
| Lens (contra) | • | • | • | • | • | • | $D_{\text{max}} < 1000$ | • | • | • |
| Eye (ipsi) | • | • | • | • | • | • | $D_{\text{max}} < 5000$ | • | • | • |
| Eye (contra) | • | • | • | • | • | • | $D_{\text{max}} < 5000$ | • | • | • |
| Optic nerve (ipsi) | • | • | • | • | • | • | $D_{0.01\text{cm}^3} < 5400$ | • | • | • |
| Optic nerve (contra) | • | • | • | • | • | • | $D_{0.01\text{cm}^3} < 5400$ | • | • | • |
| Retina | • | • | • | • | • | • | $D_{\text{max}} < 4000$ | • | • | • |
| Optic chiasm | • | • | • | • | • | • | $D_{0.01\text{cm}^3} < 5400$ | • | • | • |

## Appendix F

**Figure F.1:**
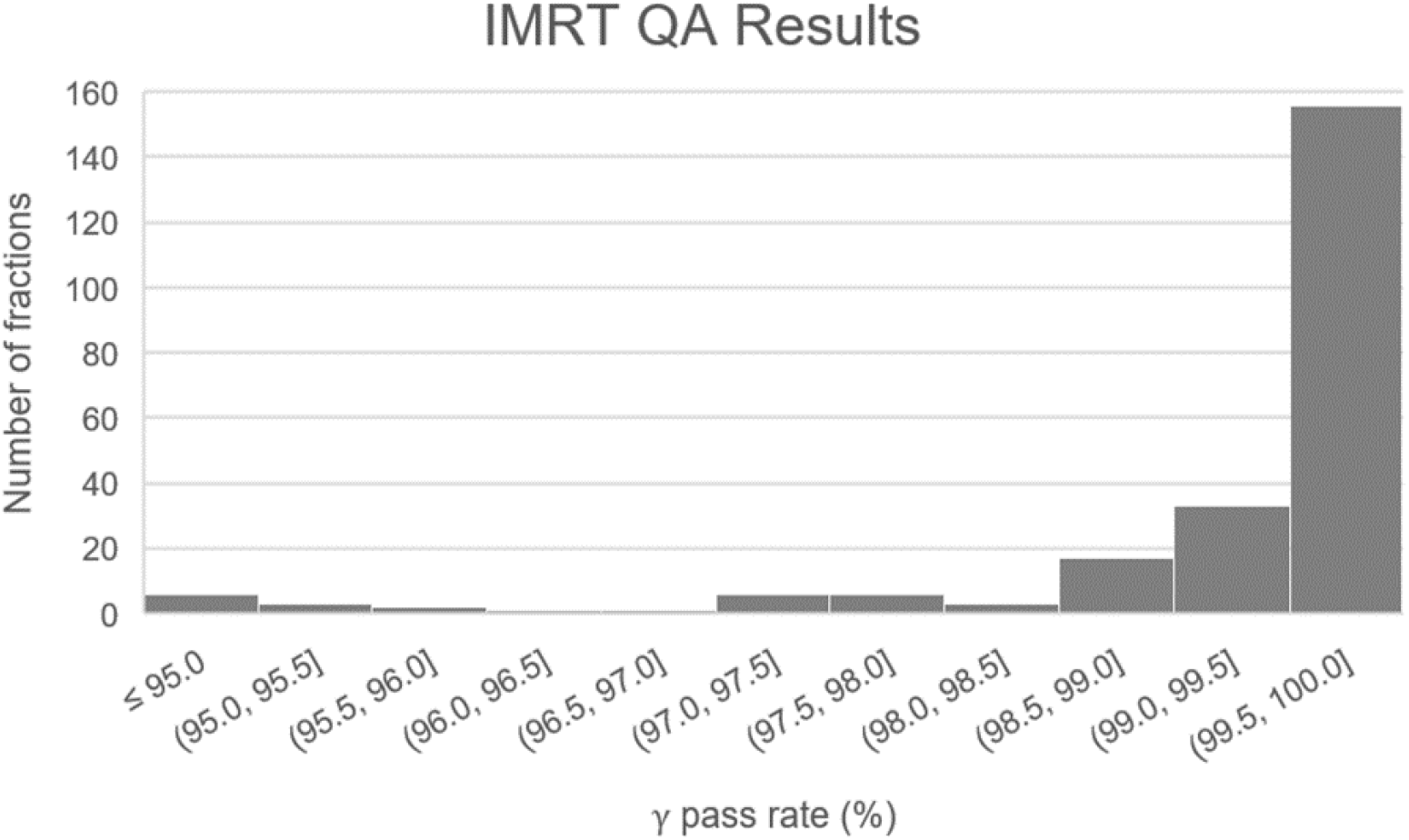
Frequency distribution of IMRT QA results for daily adaptive treatment plans. Minimum, lower quartile, median, upper quartile, and maximum are 90.9%, 99.4%, 99.9%, 100.0%, and 100.0%, respectively.

**Figure F.2:**
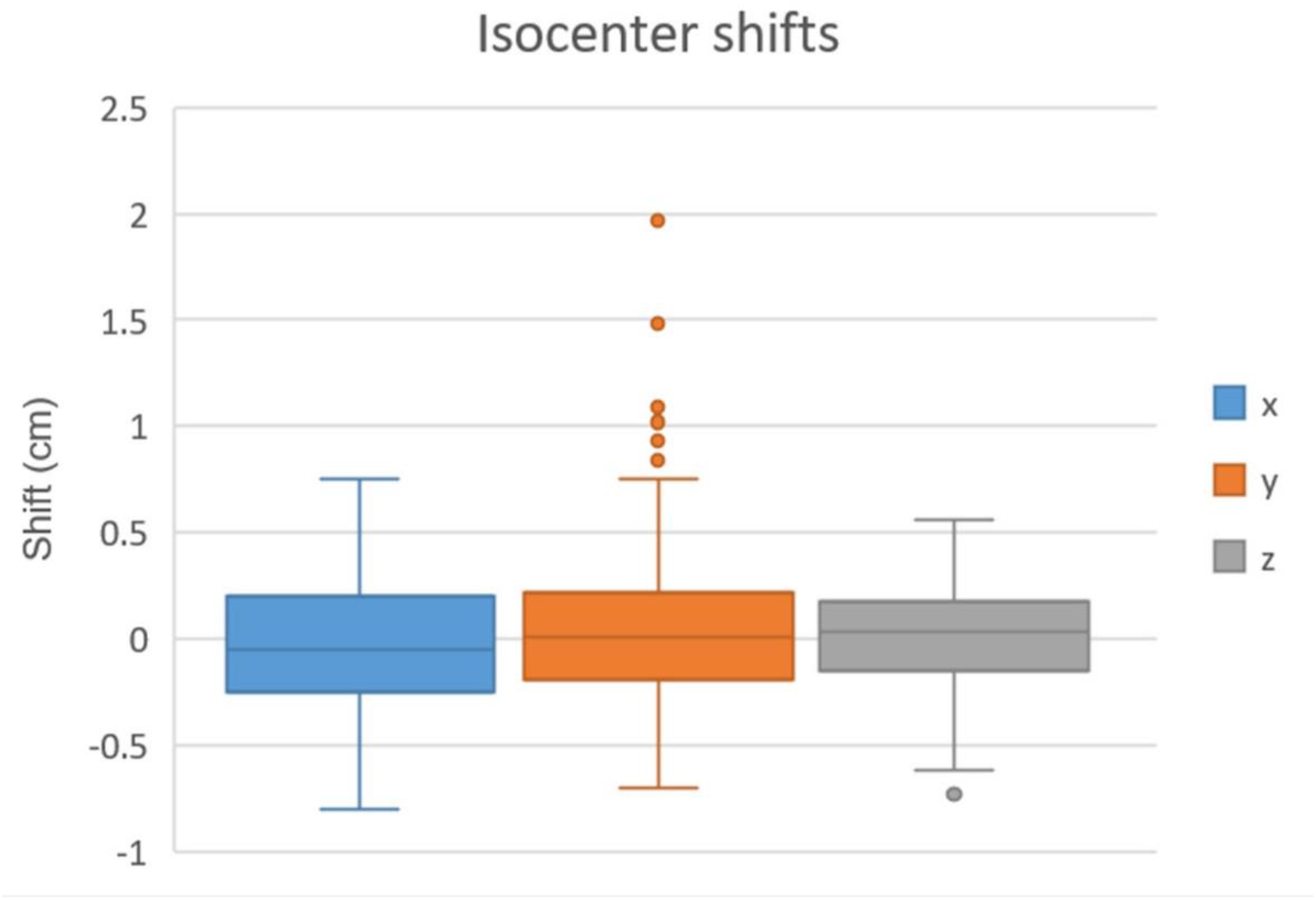
Isocenter shifts in the x (left/right), y (superior/inferior), and z (anterior/posterior) directions over 253 adaptive fractions.

## Footnotes

1 Definition of ART_ex_aequo_: “Serial plan verification to ensure pre-therapy plan parameters are stable… The new plan has the same constraints for tumor and OAR dose as the original and only allows for limited target volume adaptations and associated additional sparing of OAR, nor additional dose to the tumor.” [14]

## References

[1] Raaymakers BW, Lagendijk JJW, Overweg J, Kok JGM, Raaijmakers AJE, Kerkhof EM, et al. Integrating a 1.5 T MRI scanner with a 6 MV accelerator: Proof of concept. Physics in Medicine and Biology 2009;54:N229–37. https://doi.org/10.1088/0031-9155/54/12/N01.

[2] Lagendijk JJW, Raaymakers BW, van Vulpen M. The Magnetic Resonance Imaging-Linac System. Seminars in Radiation Oncology 2014;24:207–9. https://doi.org/10.1016/j.semradonc.2014.02.009.

[3] Winkel D, Bol GH, Kroon PS, van Asselen B, Hackett SS, Werensteijn-Honingh AM, et al. Adaptive radiotherapy: The Elekta Unity MR-linac concept. Clinical and Translational Radiation Oncology 2019;18:54–9. https://doi.org/10.1016/j.ctro.2019.04.001.

[4] Schmidt MA, Payne GS. Radiotherapy planning using MRI. Physics in Medicine and Biology 2015;60:R323–61. https://doi.org/10.1088/0031-9155/60/22/R323.

[5] Owrangi AM, Greer PB, Glide-Hurst CK. MRI-only treatment planning: Benefits and challenges. Physics in Medicine and Biology 2018;63:05TR01. https://doi.org/10.1088/1361-6560/aaaca4.

[6] Raaymakers BW, Jürgenliemk-Schulz IM, Bol GH, Glitzner M, Kotte ANTJ, van Asselen B, et al. First patients treated with a 1.5 T MRI-Linac: Clinical proof of concept of a high-precision, high-field MRI guided radiotherapy treatment. Physics in Medicine and Biology 2017;62:L41–50. https://doi.org/10.1088/1361-6560/aa9517.

[7] Nachbar M, Mönnich D, Boeke S, Gani C, Weidner N, Heinrich V, et al. Partial breast irradiation with the 1.5 T MR-Linac: First patient treatment and analysis of electron return and stream effects. Radiotherapy and Oncology 2020;145:30–5. https://doi.org/10.1016/j.radonc.2019.11.025.

[8] Bertelsen AS, Schytte T, Møller PK, Mahmood F, Riis HL, Gottlieb KL, et al. First clinical experiences with a high field 1.5 T MR linac. Acta Oncologica 2019;58:1352–7. https://doi.org/10.1080/0284186X.2019.1627417.

[9] Werensteijn-Honingh AM, Kroon PS, Winkel D, Aalbers EM, van Asselen B, Bol GH, et al. Feasibility of stereotactic radiotherapy using a 1.5?T MR-linac: Multi-fraction treatment of pelvic lymph node oligometastases. Radiotherapy and Oncology 2019;134:50–4. https://doi.org/10.1016/j.radonc.2019.01.024.

[10] Kerkmeijer LGW, Fuller CD, Verkooijen HM, Verheij M, Choudhury A, Harrington KJ, et al. The MRI-Linear Accelerator Consortium: Evidence-Based Clinical Introduction of an Innovation in Radiation Oncology Connecting Researchers, Methodology, Data Collection, Quality Assurance, and Technical Development. Frontiers in Oncology 2016;6:215. https://doi.org/10.3389/fonc.2016.00215.

[11] Verkooijen HM, Kerkmeijer LGW, Fuller CD, Huddart R, Faivre-Finn C, Verheij M, et al. R-IDEAL: A framework for systematic clinical evaluation of technical innovations in radiation oncology. Frontiers in Oncology 2017;7:1–7. https://doi.org/10.3389/fonc.2017.00059.

[12] Chuter RW, Whitehurst P, Choudhury A, van Herk M, McWilliam A. Technical Note: Investigating the impact of field size on patient selection for the 1.5T MR-Linac. Medical Physics 2017;44:5667–71. https://doi.org/10.1002/mp.12557.

[13] Kiser K, Meheissen MAM, Mohamed ASR, Kamal M, Ng SP, Elhalawani H, et al. Prospective quantitative quality assurance and deformation estimation of MRI-CT image registration in simulation of head and neck radiotherapy patients. Clinical and Translational Radiation Oncology 2019;18:120–7. https://doi.org/10.1016/j.ctro.2019.04.018.

[14] Heukelom J, Fuller CD. Head and Neck Cancer Adaptive Radiation Therapy (ART): Conceptual Considerations for the Informed Clinician. Seminars in Radiation Oncology 2019;29:258–73. https://doi.org/10.1016/j.semradonc.2019.02.008.

[15] van Herk M. Errors and Margins in Radiotherapy. Seminars in Radiation Oncology 2004;14:52–64. https://doi.org/10.1053/j.semradonc.2003.10.003.

[16] Castelli J, Simon A, Lafond C, Perichon N, Rigaud B, Chajon E, et al. Adaptive radiotherapy for head and neck cancer. Acta Oncologica 2018;57:1284–92. https://doi.org/10.1080/0284186X.2018.1505053.

[17] Bahl A, Elangovan A, Dracham CB, Kaur S, Oinam AS, Trivedi G, et al. Analysis of volumetric and dosimetric changes in mid treatment CT scan in carcinoma nasopharynx: implications for adaptive radiotherapy. Journal of Experimental Therapeutics & Oncology 2018;13:33–9.

[18] Burela N, Soni TP, Patni N, Natarajan T. Adaptive intensity-modulated radiotherapy in head-and-neck cancer: A volumetric and dosimetric study. Journal of Cancer Research and Therapeutics 2019;15:533–8. https://doi.org/10.4103/jcrt.JCRT_594_17.

[19] Hunter KU, Schipper M, Feng FY, Lyden T, Haxer M, Murdoch-Kinch CA, et al. Toxicities affecting quality of life after chemo-IMRT of oropharyngeal cancer: Prospective study of patient-reported, observer-rated, and objective outcomes. International Journal of Radiation Oncology Biology Physics 2013;85:935–40. https://doi.org/10.1016/j.ijrobp.2012.08.030.

[20] Rosenthal DI, Mendoza TR, Fuller CD, Hutcheson KA, Wang XS, Hanna EY, et al. Patterns of symptom burden during radiotherapy or concurrent chemoradiotherapy for head and neck cancer: A prospective analysis using the University of Texas MD Anderson Cancer Center Symptom Inventory-Head and Neck Module. Cancer 2014;120:1975–84. https://doi.org/10.1002/cncr.28672.

[21] Castelli J, Simon A, Louvel G, Henry O, Chajon E, Nassef M, et al. Impact of head and neck cancer adaptive radiotherapy to spare the parotid glands and decrease the risk of xerostomia. Radiation Oncology 2015;10:6. https://doi.org/10.1186/s13014-014-0318-z.

[22] Schwartz DL, Garden AS, Shah SJ, Chronowski G, Sejpal S, Rosenthal DI, et al. Adaptive radiotherapy for head and neck cancer - Dosimetric results from a prospective clinical trial. Radiotherapy and Oncology 2013;106:80–4. https://doi.org/10.1016/j.radonc.2012.10.010.

[23] Surucu M, Shah KK, Roeske JC, Choi M, Small W, Emami B. Adaptive Radiotherapy for Head and Neck Cancer: Implications for Clinical and Dosimetry Outcomes. Technology in Cancer Research and Treatment 2017;16:218–23. https://doi.org/10.1177/1533034616662165.

[24] Belshaw L, Agnew CE, Irvine DM, Rooney KP, McGarry CK. Adaptive radiotherapy for head and neck cancer reduces the requirement for rescans during treatment due to spinal cord dose. Radiation Oncology 2019;14:1–7. https://doi.org/10.1186/s13014-019-1400-3.

[25] Heukelom J, Kantor ME, Mohamed ASR, Elhalawani H, Kocak-Uzel E, Lin T, et al. Differences between planned and delivered dose for head and neck cancer, and their consequences for normal tissue complication probability and treatment adaptation. Radiotherapy and Oncology 2019. https://doi.org/10.1016/j.radonc.2019.07.034.

[26] Mohamed ASR, Bahig H, Aristophanous M, Blanchard P, Kamal M, Ding Y, et al. Prospective in silico study of the feasibility and dosimetric advantages of MRI-guided dose adaptation for human papillomavirus positive oropharyngeal cancer patients compared with standard IMRT. Clinical and Translational Radiation Oncology 2018;11:11–8. https://doi.org/10.1016/j.ctro.2018.04.005.

[27] McCulloch MM, Lee C, Rosen BS, Kamp JD, Lockhart CM, Lee JY, et al. Predictive Models to Determine Clinically Relevant Deviations in Delivered Dose for Head and Neck Cancer. Practical Radiation Oncology 2019;9:e422–31. https://doi.org/10.1016/j.prro.2019.02.014.

[28] Bahig H, Yuan Y, Mohamed ASR, Brock KK, Ping Ng S, Wang J, et al. Magnetic Resonance-based Response Assessment and Dose Adaptation in Human Papilloma Virus Positive Tumors of the Oropharynx Treated with Radiotherapy (MR-ADAPTOR): An R-IDEAL Stage 2a-2b/ Bayesian Phase II Trial. Clinical and Translational Radiation Oncology 2018;13:19–23. https://doi.org/10.1016/j.ctro.2018.08.003.

[29] Howlin C, O’Shea E, Dunne M, Mullaney L, McGarry M, Clayton-Lea A, et al. A randomized controlled trial comparing customized versus standard headrests for head and neck radiotherapy immobilization in terms of set-up errors, patient comfort and staff satisfaction (ICORG 08-09). Radiography 2015;21:74–83. https://doi.org/10.1016/j.radi.2014.07.009.

[30] Contesini M, Guberti M, Saccani R, Braglia L, Iotti C, Botti A, et al. Setup errors in patients with head-neck cancer (HNC), treated using the Intensity Modulated Radiation Therapy (IMRT) technique: How it influences the customised immobilisation systems, patient’s pain and anxiety. Radiation Oncology 2017;12:1–7. https://doi.org/10.1186/s13014-017-0807-y.

[31] Ding Y, Mohamed ASR, Yang J, Colen RR, Frank SJ, Wang J, et al. Prospective observer and software-based assessment of magnetic resonance imaging quality in head and neck cancer: Should standard positioning and immobilization be required for radiation therapy applications? Practical Radiation Oncology 2015;5:e299–308. https://doi.org/10.1016/j.prro.2014.11.003.

[32] Tijssen RHN, Philippens MEP, Paulson ES, Glitzner M, Chugh B, Wetscherek A, et al. MRI commissioning of 1.5T MR-linac systems – a multi-institutional study. Radiotherapy and Oncology 2019;132:114–20. https://doi.org/10.1016/j.radonc.2018.12.011.

[33] Kooreman ES, van Houdt PJ, Nowee ME, van Pelt VWJ, Tijssen RHN, Paulson ES, et al. Feasibility and accuracy of quantitative imaging on a 1.5 T MR-linear accelerator. Radiotherapy and Oncology 2019;133:156–62. https://doi.org/10.1016/j.radonc.2019.01.011.

## References

[1] Mehanna H, Robinson M, Hartley A, Kong A, Foran B, Fulton-Lieuw T, et al. Radiotherapy plus cisplatin or cetuximab in low-risk human papillomavirus-positive oropharyngeal cancer (De-ESCALaTE HPV): an open-label randomised controlled phase 3 trial. The Lancet 2019;393:51–60. https://doi.org/10.1016/S0140-6736(18)32752-1.

[2] Gillison ML, Trotti AM, Harris J, Eisbruch A, Harari PM, Adelstein DJ, et al. Radiotherapy plus cetuximab or cisplatin in human papillomavirus-positive oropharyngeal cancer (NRG Oncology RTOG 1016): a randomised, multicentre, non-inferiority trial. The Lancet 2019;393:40–50. https://doi.org/10.1016/S0140-6736(18)32779-X.

[3] Bahig H, Yuan Y, Mohamed ASR, Brock KK, Ping Ng S, Wang J, et al. Magnetic Resonance-based Response Assessment and Dose Adaptation in Human Papilloma Virus Positive Tumors of the Oropharynx Treated with Radiotherapy (MR-ADAPTOR): An R-IDEAL Stage 2a-2b/ Bayesian Phase II Trial. Clinical and Translational Radiation Oncology 2018;13:19–23. https://doi.org/10.1016/j.ctro.2018.08.003.

[4] Brouwer CL, Steenbakkers RJHM, Bourhis J, Budach W, Grau C, Grégoire V, et al. CT-based delineation of organs at risk in the head and neck region: DAHANCA, EORTC, GORTEC, HKNPCSG, NCIC CTG, NCRI, NRG Oncology and TROG consensus guidelines. Radiotherapy and Oncology 2015;117:83–90. https://doi.org/10.1016/j.radonc.2015.07.041.

[5] Grégoire V, Evans M, Le QT, Bourhis J, Budach V, Chen A, et al. Delineation of the primary tumour Clinical Target Volumes (CTV-P) in laryngeal, hypopharyngeal, oropharyngeal and oral cavity squamous cell carcinoma: AIRO, CACA, DAHANCA, EORTC, GEORCC, GORTEC, HKNPCSG, HNCIG, IAG-KHT, LPRHHT, NCIC CTG, NCRI, NRG Oncolog. Radiotherapy and Oncology 2018;126:3–24. https://doi.org/10.1016/j.radonc.2017.10.016.

[6] Grégoire V, Ang K, Budach W, Grau C, Hamoir M, Langendijk JA, et al. Delineation of the neck node levels for head and neck tumors: A 2013 update. DAHANCA, EORTC, HKNPCSG, NCIC CTG, NCRI, RTOG, TROG consensus guidelines. Radiotherapy and Oncology 2014;110:172–81. https://doi.org/10.1016/j.radonc.2013.10.010.

[7] van Herk M, Remeijer P, Rasch C, Lebesque J v. The probability of correct target dosage: Dose-population histograms for deriving treatment margins in radiotherapy. International Journal of Radiation Oncology Biology Physics 2000;47:1121–35. https://doi.org/10.1016/S0360-3016(00)00518-6.

[8] McKenzie A, van Herk M, Mijnheer B. Margins for geometric uncertainty around organs at risk in radiotherapy. Radiotherapy and Oncology 2002;62:299–307. https://doi.org/10.1016/S0167-8140(02)00015-4.

[9] Yang J, Garden AS, Zhang Y, Zhang L, Dong L. Variable planning margin approach to account for locoregional variations in setup uncertainties. Medical Physics 2012;39:5136–44. https://doi.org/10.1118/1.4737891.

[10] Wang H, Wang C, Tung S, Dimmitt AW, Wong PF, Edson MA, et al. Improved setup and positioning accuracy using a three-point customized cushion/mask/bite-block immobilization system for stereotactic reirradiation of head and neck cancer. Journal of Applied Clinical Medical Physics 2016;17:180–9. https://doi.org/10.1120/jacmp.v17i3.6038.

[11] Ding Y, Mohamed ASR, Yang J, Colen RR, Frank SJ, Wang J, et al. Prospective observer and software-based assessment of magnetic resonance imaging quality in head and neck cancer: Should standard positioning and immobilization be required for radiation therapy applications? Practical Radiation Oncology 2015;5:e299–308. https://doi.org/10.1016/j.prro.2014.11.003.

[12] Kiser K, Meheissen MAM, Mohamed ASR, Kamal M, Ng SP, Elhalawani H, et al. Prospective quantitative quality assurance and deformation estimation of MRI-CT image registration in simulation of head and neck radiotherapy patients. Clinical and Translational Radiation Oncology 2019;18:120–7. https://doi.org/10.1016/j.ctro.2019.04.018.

